# Microbiological Quality of Weaning Foods and Hygiene-Related Risk Factors in Peri-Urban, Lusaka Zambia

**DOI:** 10.64898/2026.06.23.26356408

**Authors:** Jenala Chipungu, Dennis Ngosa, Sarah Bick, Katherine Davies, Katayi Mwila-Kazimbaya, Anjali Sharma, Laura Braun, Roma Chilengi, Jacqueline Knee, Robert Dreibelbis

## Abstract

Food contamination contributes to 40% of childhood diarrhoea cases globally. It occurs when pathogens are transmitted from faecal matter to food through the faecal oral route. Food hygiene can prevent food contamination and improve the microbial quality of weaning foods in domestic settings. However, context specific evidence is needed to identify risk factors associated with food contamination, especially in complex low-income communities. Our study used a modified Hazard Analysis Critical Control Point approach to assess the quality of weaning foods and identify associated risk factors in a low income setting of Lusaka, Zambia. We enrolled 60 caregivers of children aged ≤1 year who had begun complementary feeding and collected data on household characteristics and food hygiene behaviours using surveys and structured observations. We collected samples of complimentary foods prepared for the child and tested them for *Escherichia coli (E. coli)* using the IDEXX Colilert-18 method. Multivariable logistic regression was performed to determine risk factors associated with food contamination. Of 59 food samples, 17 (29%) were contaminated with *E. coli*. Porridge (AOR = 0.04; 95% CI: 0.01, 0.28; p = 0.001) and non-animal source foods (AOR = 0.11; 95% CI: 0.02, 0.69; p = 0.019) were associated with lower odds of contamination compared with animal source foods. Heating of food was also associated with lower odds of contamination but was not significant. Food hygiene behaviours including utensil and surface cleaning were seldom practiced (17% and 15%, respectively), and handwashing with soap before food preparation was not practiced. Our study identifies the microbiological risk associated with animal source foods and the potential health risk posed to weaning children. Nutrition guidelines should take into consideration this risk as programs promote an increased uptake of animal source foods in child diets.

## Introduction

Diarrhoeal disease claims 525,000 deaths in children under-5 years, with up to 90% of these deaths occurring in low-income countries (1). Contaminated food contributes to 40% of under-5 childhood diarrhoea cases globally (2). Diarrhoea episodes increase during the weaning period when children are increasingly exposed to enteric pathogens as they transition from breastfeeding to solid foods (3). Several studies in low resource settings have found that weaning foods are often heavily contaminated with faecal indicator bacteria (FIB) (4–6). The transmission pathway for FIB to food follows the faecal oral route, where FIB travels from an infected person to another via contaminated water, food, surfaces and hands (7). Food hygiene, defined as conditions and measures necessary to ensure microbial food safety, therefore becomes a key measure in reducing weaning food contamination (8).

The global food hygiene guidelines for household settings state five key measures for food safety: keep surfaces and hands clean, separate raw and cooked food, cook thoroughly, keep food at safe temperatures, and use safe water and raw materials (8). However, the application of these guidelines in low-income settings can be challenging. In sub-Saharan Africa, approximately 50% of the population have access to electricity, and only 32% and 27% have access to safe water and hand hygiene services, respectively (9, 10). These conditions hinder the refrigeration of food at safe temperatures, use of safe water during food preparation and hand washing with soap during food preparation and feeding.

Despite its important role in food safety, food hygiene in the domestic setting in low-income regions has received limited attention in research and guidelines. Child nutrition programmes have provided limited guidance on specific food hygiene behaviours. For example, the Infant and Young Child Feeding (IYCF) global initiative recommends practicing proper hygiene and food handling during food preparation but does not provide specific guidance on what hygiene behaviours are necessary to ensure microbial food quality (11). Braun et al found only 11 robust impact evaluations of food hygiene interventions eligible for inclusion in their meta-analysis (12). These interventions often targeted behaviours such as utensil care, handwashing with soap, re-heating of stored food and surface cleaning to improve the microbial quality of weaning foods and reduce child diarrhoea (13–16). However, the process of selecting which behaviours to use in interventions aimed at improving microbiological food quality require context specific evidence that indicate the risk factors for food contamination.

To build the evidence base on which behaviours to target for food hygiene interventions, we investigated potential risk factors and their associations with weaning food contamination in peri-urban Zambia. The findings from this research will provide direction as to which food hygiene behaviours are important to consider in the promotion of food hygiene interventions in domestic settings.

## Methods

Our study used a modified hazard analysis and critical control point (HACCP) approach to conduct a mixed methods cross-sectional study in Lusaka, Zambia. The HACCP approach describes specific steps in the food production industry to identify potential food hazards and their corrective measures (17). We used structured observations to document food hygiene behaviours and household surveys to characterise our population. Microbial food quality was determined by the presence or absence of *Escherichia coli (E. coli*) - a faecal indicator bacterium.

### Study Setting

The study was conducted in George, a low-income neighbourhood in Lusaka, Zambia, with a population of approximately 70,000 people. It is characterised by rapid population growth, unplanned housing, reliance on unimproved shared sanitation, absence of drainage and waste management systems, poor access to clean water and poverty (18). Households typically live on plots subdivided into an average of four houses per plot (19).

### Sample Size and Sampling

Sixty (60) caregivers, residing in George neighbourhood with a child aged 1 year or younger were recruited between the 6^th^ of June 2024 and the 4^th^ of July 2024. This sample size was determined based on similar HACCP studies conducted previously (13–16). To capture a variation of households, we used a drawn map of George and divided the neighbourhood into 4 roughly equal sections and selected 15 households from each section. The first plot was selected from a common landmark using the spin the bottle method. Within the plot, the first house was approached and enrolled if eligible. If they did not meet the eligibility criteria, the next household in the plot was approached. We approached every third plot thereafter until the total sample of 60 was reached.

Sampling and all data collection were conducted by a team of 4 enumerators (2 females and 2 males) that received 5 days of training on all data collection and sample collection procedures as well as ethical research conduct.

### Structured observations

Upon enrolment, caregivers were told that a research assistant would visit them the next day during the time they normally prepare food for their child, and the food preparation process would be observed. A structured observation guide was created to capture critical food hygiene behaviours practiced or missed from the start of the food preparation process until after the child was fed. We observed handwashing with soap before food preparation, type of food being prepared, whether food was previously cooked, the elevation of the surface used to prepare food, surface cleaning behaviours, utensil cleaning, type of cooking fuel used and whether food was heated. Observations were non-participatory, and data was entered using electronic data capture forms in Open Data Kit (ODK) on handheld tablets.

### Food sample collection

After weaning food was prepared, the research assistant used a sterile disposable 10mg spoon provided by the study to collect two spoonfuls of food and deposited it into a sterile 100ml whirl-Pak bag. Where more than one type of food was prepared, two spoonfuls of each food type was collected and placed into the same bag. Each sample was labelled with the participant ID, date of sample collection, time of collection and type of food. Samples were stored in cool bags with cold packs and delivered to the laboratory within 3hours of collection.

### Household survey

Once the observation was completed, research assistants administered a household survey and collected data on access to water and sanitation, access to cooking fuel and weaning foods. Water, sanitation and hygiene (WASH) questions included type of sanitation facility used, water source and availability of water, and presence of handwashing station with water and soap. A 4-question individual water insecurity (IWISE) validated scale was also included (20) to assess water insecurity experienced by participants. Questions on cooking fuel included type of cooking fuel used for cooking and its availability and questions on weaning food covered food storage behaviours and access to food. Socio-demographic questions were also asked and included age, marital status, education status and household wealth. Survey data was captured using Open Data Kit (ODK) and administered using handheld tablets.

### Microbiological analysis

Food contamination was determined by the presence or absence *E. coli*. The IDEXX Colilert-18® rapid culture-based method was used to detect and quantify *E. coli* in food samples. Each food sample was first manually homogenised using a masher, 15g of which was transferred to 200ml whirl Pak bag and diluted with 100ml of sterile distilled water. The bag was shaken vigorously by hand for 60 seconds and put aside to settle for 15 minutes at room temperature (between 18 and 24 degrees Celsius). 100ml of the supernatant was then transferred into a 120ml sterile sample bottle, mixed with Colilert reagent powder until fully dissolved, and sealed in a IDEXX Quanti-Tray/2000. A negative control of sterile water was processed alongside each batch of samples daily. The samples were incubated for 18hrs to 22hrs at 35 ± 0.5 °C. Yellow cells in the IDEXX Quanti tray represent presence of coliforms while fluorescent wells under UV light represent presence of *E. coli*. The number of yellow cells and fluorescent wells were enumerated and recorded. Using the Most Probable Number (MPN) IDEXX chart we estimated the *E. coli* bacterial count per 10g of sample.

### Statistical Analysis

The primary outcome was any sample with an *E. coli* count above zero. All analyses were conducted in Stata version 18.5 (Stata Corp, College Station, TX, USA). We used descriptive statistics to summarize data from microbiological assessments, surveys, and structured observations, and reported them as frequencies (n) and proportions (%). Apart from food type, all variables of interest were dichotomized. The household wealth variable was derived from the Demographic Health Survey (DHS) composite Wealth Index that assesses household assets. A principal component analysis (PCA) was performed to generate five wealth quintiles which were then dichotomised by combining quintiles 1-3 into the lowest wealth group and quintiles 4 – 5 into the upper wealth group. Food type was categorized into three groups: porridge, animal-source foods, and non–animal-source foods. The porridge category included both instant porridge and traditionally prepared maize meal porridge; animal-source foods comprised chicken, beef, pork, and eggs; and non–animal-source foods included beans, potatoes and green leafy vegetables. We also assessed the median *E. coli* count (CFU/10g) by food group using the Kruskal-Wallis test.

Variables of interest correspondent to specific risk factors (reported and observed) and based on previous HACCP studies (13–16, 21) were cross tabulated with the primary outcome. Crude associations were assessed using Fisher’s exact test. Those with a *p* value <0.20 were considered as candidate variables for a logistic regression to identify risk factors of *E. coli* contamination.

All variables that met inclusion criteria in the first step of the analysis (Fisher’s exact p-value < 0.2) were included in a multivariable logistic regression model. This was followed by an investigator-led iterative backward stepwise process with removal of the variable with the highest p-value at each step. Goodness-of-fit of the new model compared to the base model was assessed by comparing the Bayesian Information Criterion (BIC). Once all variables had been checked once, the process was repeated until a final parsimonious model was produced where no further removal improved model fit.

### Ethical Considerations

Ethical approvals were obtained from the University of Zambia Biomedical Research Ethics Committee (UNZABREC), (REF: 4403-2023) and the London School of Hygiene and Tropical Medicine (LSHTM), (REF: 29572). All participants were given detailed information on the study and an option for an impartial witness if illiterate. Written consent was obtained, and all enrolled participants were reimbursed with ZMW 150 Zambian Kwacha (approx. USD 7) for their time and for the food sample they provided. All data was anonymised prior to analysis.

## Results

One food sample did not meet quality control standards and data from that sample and associated participant record was removed from analysis. A total of 59 records were available with complete household demographics, structured observation, household survey, and microbiological outcomes.

### Household Characteristics

All participants were female (n=59); the majority were in the 25 to 34 years age group (45%), married (68%), had a family size of 2 to 5 members (53%), attained at least secondary education (53%) and were in the lower wealth group (72%). Charcoal was the main cooking fuel (97%), with 46% reporting having cooking fuel only sometimes. 90% had access to an improved water source, 56% experienced water insecurity and 90% relied on a mobile object to practice handwashing, 92% accessed weaning foods from the local market and 58% reported always covering stored food.

### Observed Food Hygiene Behaviours

There was no handwashing with soap observed before food preparation (Table 2). Most food was prepared on the floor (84%) and surface and utensil cleaning behaviours before use were minimally observed (15% and 17%, respectively). 73% of meals were freshly cooked and 83% were heated during the food preparation process.

### Food Microbiological contamination

Analysed food samples were eggs (n=7), beans (n=4), meat including beef, chicken or sausage (n=6), vegetables (n=15), and porridge (n=27), (Figure 1). Apart from porridge, all food samples were mainly served with nshima (a traditional staple food made from maize meal and cooked to a firm dough-like consistency). Of the 59 food samples, 17 (29%) tested positive for *E. coli* (Table 2). Animal source foods had the highest proportion of contaminated samples (69%) compared with non-animal source foods (32%) and porridge (7%) (Fisher’s exact p-value <0.001). The median *E. coli* count was 4.1 CFU/10g for animal source foods compared with zero for non-animal source foods and porridge.

**Figure 1:**
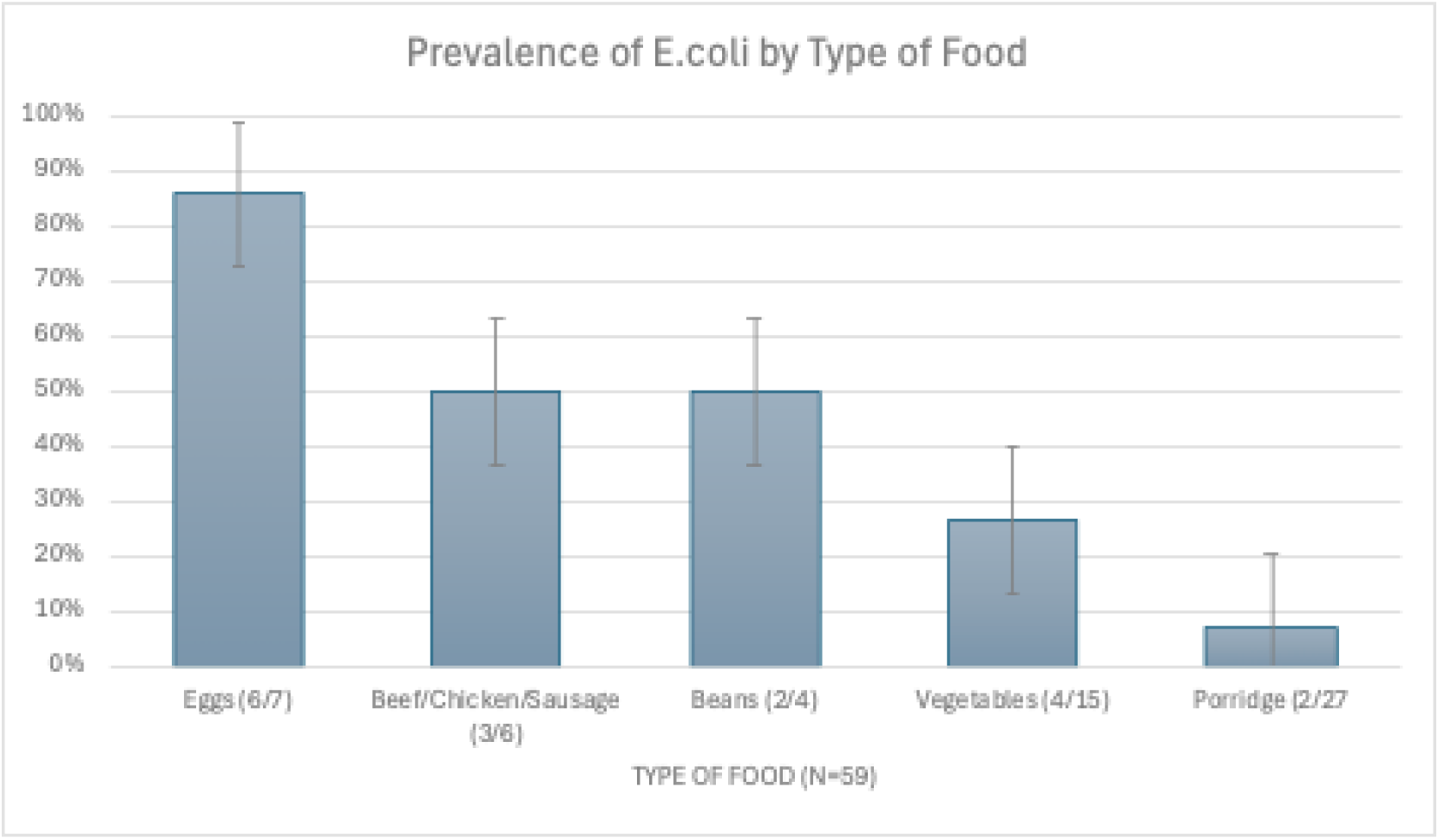

### Proportion of food samples contaminated with E. coli

Contaminated samples were highest in the upper wealth group (44% vs 23%, p = 0.19), those with intermittent cooking fuel availability (41% vs 19%, p = 0.086), previously cooked foods (50% vs 21%, *p* = 0.05) and foods that were not heated (60% vs 24% p = 0.054), (Table 1 and Table 2). Along with food type, these variables met the inclusion criteria for multivariable logistic models.

**Table 1:**
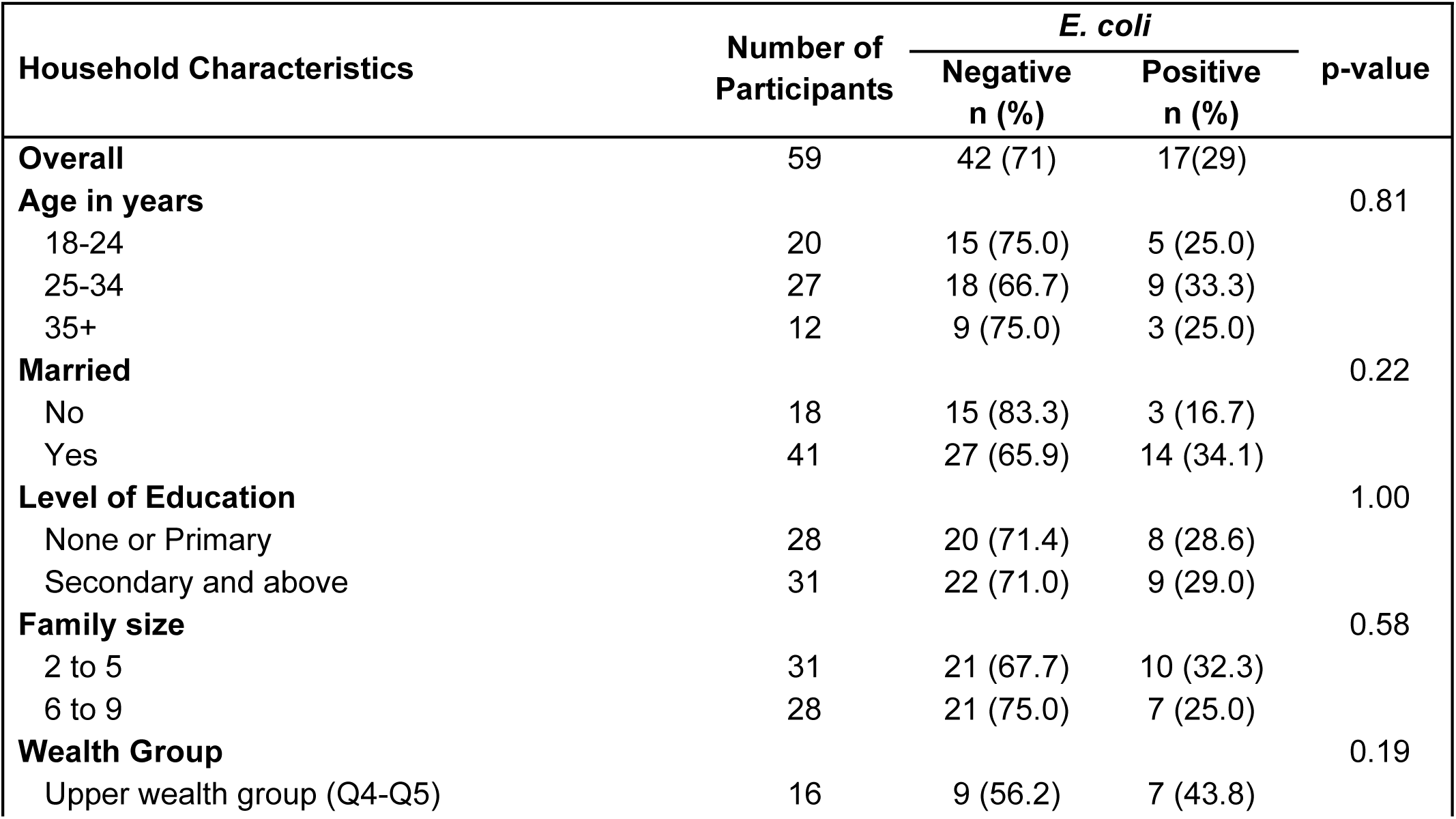
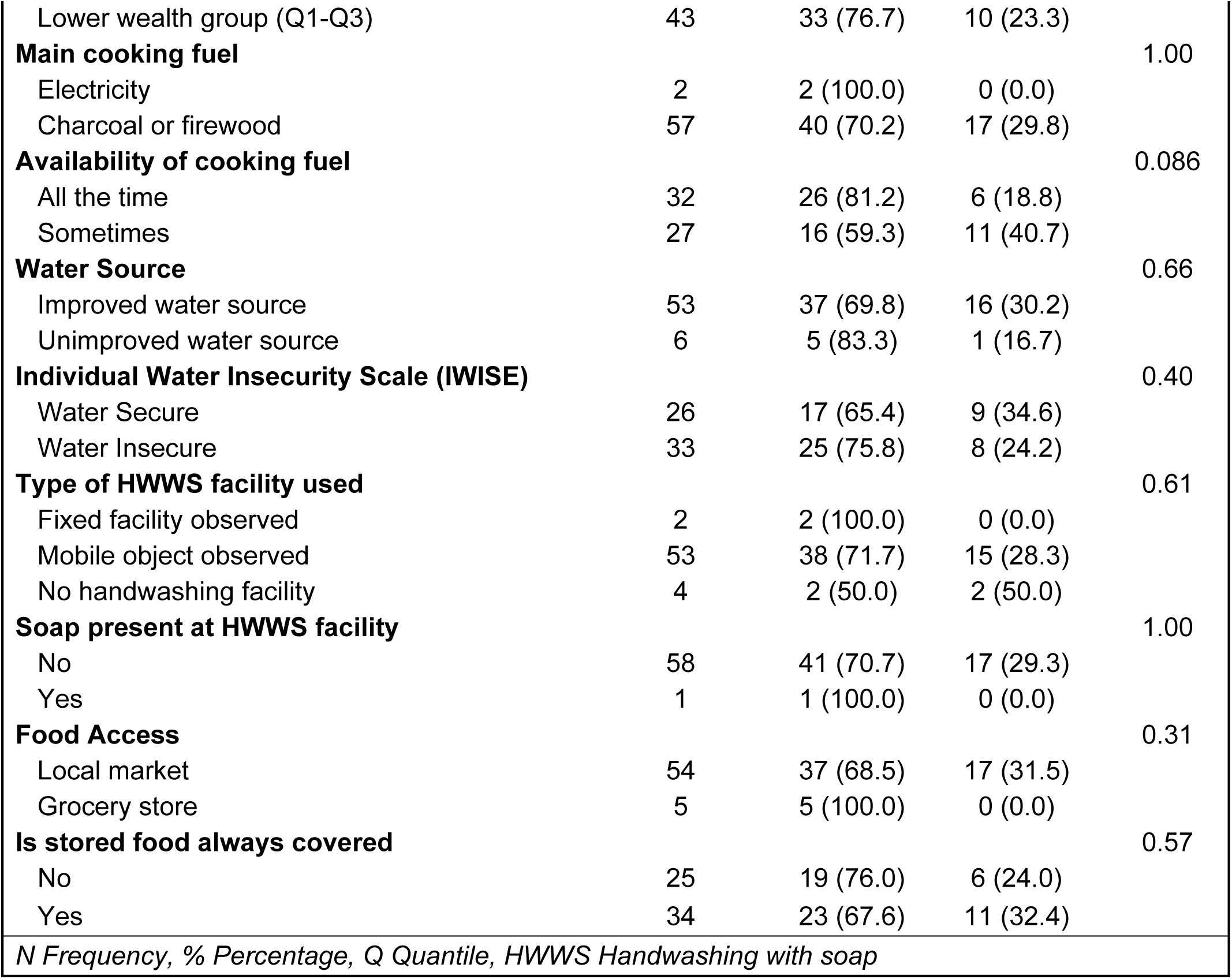
**Household Characteristics by Presence of *E. coli***

**Table 2:**
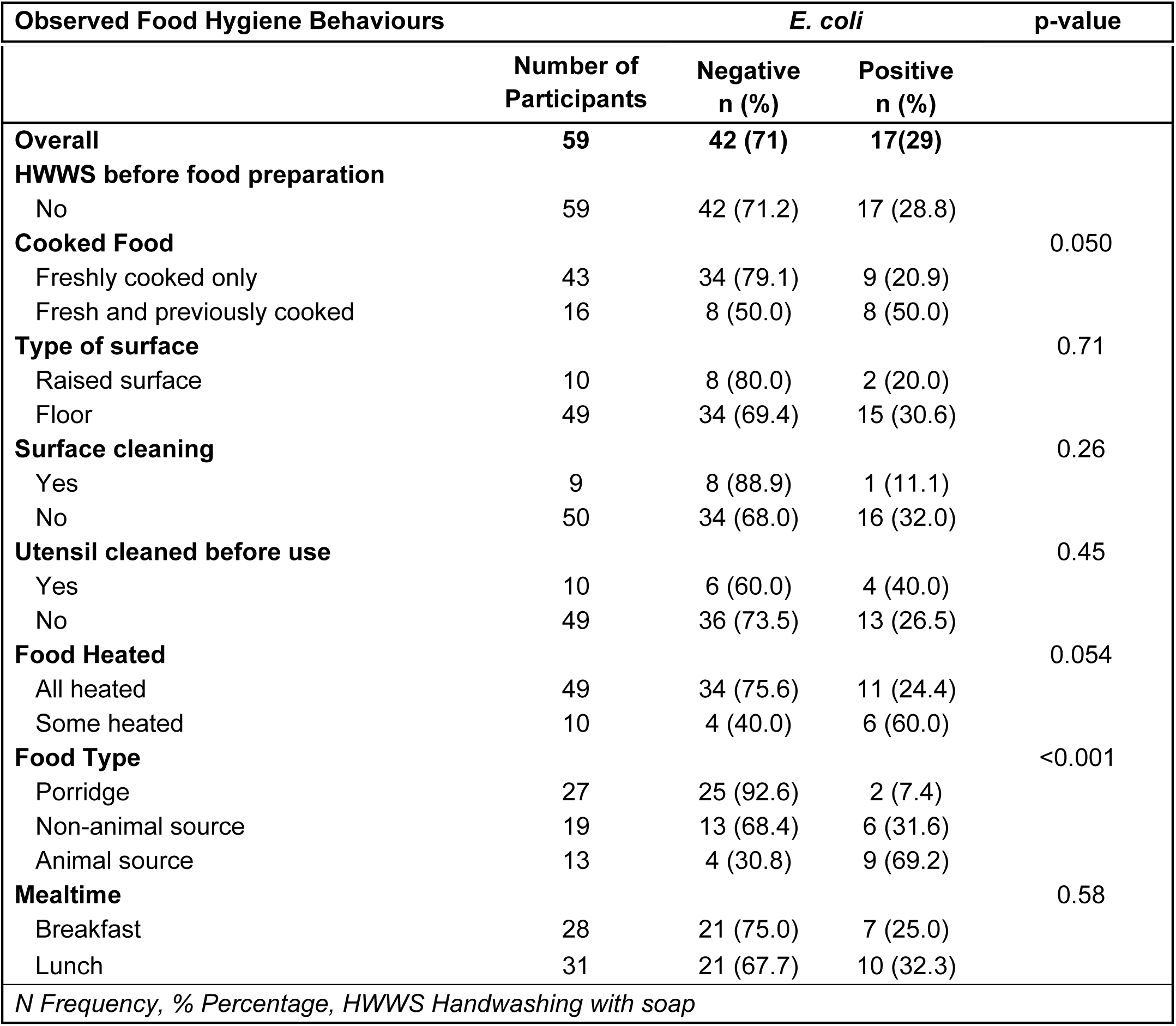
**Observed Food Hygiene Behaviours by Presence of *E. coli***

| Observed Food Hygiene Behaviours | *E. coli* | p-value |
| --- | --- | --- |

|  | Number of<br>Participants | Negative<br>n (%) | Positive<br>n (%) |  |
| --- | --- | --- | --- | --- |
| **Overall** | **59** | **42 (71)** | **17(29)** |  |
| **HWWS before food preparation** |  |  |  |  |
| No | 59 | 42 (71.2) | 17 (28.8) |  |
| **Cooked Food** |  |  |  | 0.050 |
| Freshly cooked only | 43 | 34 (79.1) | 9 (20.9) |  |
| Fresh and previously cooked | 16 | 8 (50.0) | 8 (50.0) |  |
| **Type of surface** |  |  |  | 0.71 |
| Raised surface | 10 | 8 (80.0) | 2 (20.0) |  |
| Floor | 49 | 34 (69.4) | 15 (30.6) |  |
| **Surface cleaning** |  |  |  | 0.26 |
| Yes | 9 | 8 (88.9) | 1 (11.1) |  |
| No | 50 | 34 (68.0) | 16 (32.0) |  |
| **Utensil cleaned before use** |  |  |  | 0.45 |
| Yes | 10 | 6 (60.0) | 4 (40.0) |  |
| No | 49 | 36 (73.5) | 13 (26.5) |  |
| **Food Heated** |  |  |  | 0.054 |
| All heated | 49 | 34 (75.6) | 11 (24.4) |  |
| Some heated | 10 | 4 (40.0) | 6 (60.0) |  |
| **Food Type** |  |  |  | <0.001 |
| Porridge | 27 | 25 (92.6) | 2 (7.4) |  |
| Non-animal source | 19 | 13 (68.4) | 6 (31.6) |  |
| Animal source | 13 | 4 (30.8) | 9 (69.2) |  |
| **Mealtime** |  |  |  | 0.58 |
| Breakfast | 28 | 21 (75.0) | 7 (25.0) |  |
| Lunch | 31 | 21 (67.7) | 10 (32.3) |  |
| *N Frequency, % Percentage, HWWS Handwashing with soap* |  |  |  |  |

### Risk Factors of Food Microbiological Contamination

In the multivariable logistic regression analysis, food heating, porridge (Odds Ratio (OR) = 0.03; 95% CI: 0.00, 0.25; p = 0.001) and non-animal source food (OR = 0.10; 95% CI: 0.01, 0.67; p = 0.018) were associated with lower odds of food contamination (Table 3). In the backwards selection process, cooked food, cooking fuel availability, and wealth variables were consecutively removed from the model based on their high p-values. The final parsimonious model comprised of food heating and food type, with porridge (Adjusted Odds Ratio (aOR) = 0.04; 95% CI: 0.01, 0.28; p = 0.001) and non-animal source foods (aOR = 0.11; 95% CI: 0.02, 0.69; p = 0.019) being significantly associated with lower odds of contamination compared with animal source foods.

**Table 3:** Multivariable Logistic Regression: Risk Factors Associated with Food Contamination.

| Risk Factors | Adjusted<br>(Full Model) |  | Adjusted<br>(Parsimonious Model) |  |
| --- | --- | --- | --- | --- |
|  | OR (95% CI) | p-value | OR (95% CI) | p-value |
| **Wealth group** |  |  |  |  |
| Lower wealth group (Q1 - Q3) | Ref | - |  |  |
| Upper wealth group (Q4 and Q5) | 3.35 (0.66, 17.07) | 0.145 |  |  |
| **Cooking Fuel Availability** |  |  |  |  |
| Sometimes | Ref | - |  |  |
| All the time | 0.40 (0.09, 1.90) | 0.240 |  |  |
| **Cooked Food** |  |  |  |  |
| Fresh and previously cooked | Ref | - |  |  |
| Freshly cooked | 3.77 (0.27, 52.8) | 0.325 |  |  |
| **Food heated** |  |  |  |  |
| Some heated | Ref | - | Ref |  |
| All heated | 0.08 (0.00, 1.46) | 0.088 | 0.18 (0.03, 1.27) | 0.086 |
| **Food Type** |  |  |  |  |
| Animal source | Ref | - | Ref | - |
| Porridge | 0.03 (0.00, 0.25) | 0.001 | 0.04 (0.01, 0.28) | 0.001 |
| Non-animal source | 0.10 (0.01, 0.67) | 0.018 | 0.11 (0.02, 0.69) | 0.019 |
| OR Odds ratio, % Percentage, CI Confidence Interval, Ref Reference, Q Quantile |  |  |  |  |

## DISCUSSION

In this mixed method exploratory study using a modified HACCP approach, we investigated the quality of weaning foods and their associated risk factors among caregivers living in a peri-urban area of Lusaka, Zambia. Findings showed 29% of food samples in our study were contaminated with *E. coli*. Non-animal source foods and porridge were less frequently contaminated than animal-source foods. Direct observations show low rates of handwashing with soap and surface and utensil cleaning behaviours that could pose additional risk to the microbial quality of food in this context.

The association between food type and microbial food quality identified in this study and others, (22, 23, 5) expands the scope of food hygiene beyond mere practice of hygiene behaviours. Animal source foods are a rich source of nutrients for children and diets enriched with animal source food can avert malnutrition and stunting (24). However, in our study, they had the highest prevalence of contamination with *E. coli*. Animal source foods like meats, eggs and milk are wet foods and provide ideal conditions for the rapid growth of FIB (25). They are also more likely to become contaminated during production and supply, especially in low-income settings where meats are sold in open markets and often rinsed with contaminated water (26). Nutrition programmes recommend that animal source foods should be introduced from 6months and frequently fed for the optimal growth of children (27). However, an increased intake of animal source foods poses a potential risk to child health as they are at a higher risk of being contaminated. In view of our study findings, guidelines on child nutrition should be informed by evidence on how to best to handle and prepare animal source foods to control or reduce FIB contamination levels.

Heating of food was protective against FIB contamination, and these findings are consistent with other HACCP studies conducted in similar settings (4, 6). Food heated up to 70 degrees Celsius is enough to eliminate food borne pathogens and make weaning food safe for consumption (8). Almost all our participants relied heavily on charcoal for cooking fuel. However, a heavy reliance on charcoal and basic traditional stoves, is time consuming and can hinder adequate heating of food, especially where households cannot afford a sufficient supply. Interventions promoting the use of modern cooking stoves like liquid petroleum gas cookers have recorded improvements in child nutrition and food security (28, 29). Other factors such as low knowledge levels on the role of temperature in eliminating faecal contamination can also lead to poor food heating practices (30). Additionally, not all food types can be heated to high boiling temperatures and others are prepared by frying. Studies promoting heating for improved microbial food quality, should explore these dynamics and establish how best they can be implemented in low-income communities.

Water related hygiene behaviours including, surface and utensil cleaning were infrequently observed, and handwashing with soap was not practiced at all before food preparation. Barriers to handwashing with soap at the individual (motivation, knowledge), social (norms), structural and institutional levels have been widely documented (31–33). However, there is need to identify the opportunities that arise for handwashing with soap and other cleaning behaviours during food preparation and feeding. This is particularly important in cases where food can become contaminated by hands at the beginning or during food preparation, but contamination is later mitigated by high cooking temperature at the end of food preparation. Some HACCP studies have documented food preparation flow charts around this process and suggest hand washing with soap is practiced right before feeding or eating and utensil cleaning is practiced before serving food (5, 16). Such information is important to provide clear messaging on what food hygiene behaviours to promote and when they should be practiced during the food preparation and feeding key juncture. A better understanding of this will contribute to more precise measurement of hygiene practices and their associations with microbial food quality in the domestic setting.

## Limitations

Our study had several limitations. The relatively small sample size limited the statistical power to detect significant associations, specifically for variables with low prevalence like observed hygiene behaviours. Second, we did not collect an objective measure for food temperature limiting our ability to report whether heating of food was above or below the required thresholds. Third, our food sample comprised a combination of nshima with animal source food or a non-animal source food, which made it impossible to assess the microbial quality of the nshima or other individual food components. Consequently, the findings presented should be interpreted with caution and additional studies with larger sample sizes and more comprehensive food sampling are recommended to strengthen this evidence.

## Conclusion

The weaning period exposes children to faecal pathogens via food contamination in low-income settings. Our study highlights the potentially under-appreciated microbiological risk associated with animal sourced foods for infants, and nutrition programmes should consider the potential exposure risks these foods bring. Other protective factors were investigated, yet food hygiene remains challenging in these complex environments. Parallel ethnographic research informed by this study will further explore the challenges underlying food hygiene behaviours and provide context specific evidence to inform intervention strategies. Finally, given the persistent public health threat of childhood diarrhoea in Zambia and neighbouring countries, further evidence on food hygiene and the microbiological quality of weaning foods are needed.

## Data Availability

The minimal data set will be uploaded at LSHTM Data Compass repository and will be made available via a DOI hyperlink that is yet to be created

## Acknowledgments

We would like to thank the research assistants, namely, Kabwe Mwamba, Esther Hamweemba, Mainza Syulikwa and Palicha Halwiindi, who collected the data. We also thank the neighbourhood health committee members under George Health Centre for their support in the field and the participants who volunteered to participate in the study.

